# Assessing the Adequacy of Health Facilities for the Aging Population in Indian Cities

**DOI:** 10.1101/2024.03.15.24304353

**Authors:** A.H. Sruthi Anil Kumar, Nawaj Sarif, Papai Barman

## Abstract

**Objectives:** The healthcare system in India is struggling to meet the needs of the current population, and the rising aging population is likely to exacerbate this problem. Therefore, this study tried to investigate the distribution of the older population’s access to improved health facilities in Indian cities.

**Methods:** The study uses data from the 2011 Census of India, which is the latest census round. An index was developed to understand the available health facilities and a geospatial approach was adopted to understand the aging pattern and access to health facilities across Indian cities.

**Results:** Despite similar aging scenarios, health infrastructure was not equally distributed across cities. The study revealed that advanced health facilities were more concentrated in specific pockets, particularly in metropolitan cities, while smaller cities lacked good health facilities and were not easily accessible.

**Discussion:** The results highlight the infrastructure gap in Indian cities which poses a significant challenge in achieving healthy aging.

## 1. Introduction

Rapid reductions in fertility and mortality have resulted in global population aging (United Nations 2019; 2022). Population aging is characterized by the rise in the proportion of older persons aged 60 and above (internationally, ‘older persons’ is defined as those aged 65 and above; in India, due to the long prevailing relatively lower life expectancy at birth, older persons are defined as those aged 60 and above). At the global level, the total fertility rate (TFR) declined from 5.0 during 1950-55 to 2.3 in 2021, which is projected to reach 2.1 by 2050 (United Nations, 2022). Along with this, the achievements in mortality reduction have been significant; life expectancy at birth increased from 47 years during 1950-55 to 72.8 years in 2019, a gain of 25.8 years. Life expectancy at birth is further expected to reach 77.2 years by 2050. These two phenomena of decreased fertility and increased life expectancy have resulted in a demographic dividend and, more importantly, in population aging across nations (United Nations, 2022; 2019). However, the intercountry differences in levels and pace of fertility and mortality decline have resulted in differences in the levels and trends in population aging (United Nations, 2017).

As per World Population Aging Report, 2020, 9.3 percent of the global population (the world population size was 7.75 billion in 2020 and is expected to reach 8 billion by 15 November 2022) will be those aged 65 years and above, thus accounting for approximately 745 million older persons across the globe (United Nations, 2020). Projections indicate that this share will increase to 16 percent by 2050. Data from around the world and population projections indicate that a great extent of the increase in the absolute number of older persons aged 60 or 65 and above will be in developing nations (United Nations, 2020).

## 2. Aging in India

India has experienced significant changes in fertility and mortality in recent decades. The average number of children per woman (TFR) in India decreased from 5.9 in 1961 to 2.0 during 2019-21 (Rele, 1987; International Institute for Population Sciences, 2020), a decline of 3.9 children per woman. This fertility change occurredwith remarkable improvements in mortality; life expectancy at birth in India increased from 42.0 years in 1961 to 69.7 years during 2015-2019, an increase of 27.7 years during the interim period (Technical Group on Population Projections, 2019; United Nations, 2019). These changes have resulted in India experiencing a demographic dividend with vast interstate differences. Along with this, the states have been experiencing an increase in the number of older persons aged 60 and above at different paces.

In India, according to the latest census of 2011, the proportion aged 60 and above is 8.3 percent of the total population or 98.5 million persons. Population projections show that the share of older persons will be 11.5 percent (163.2 million) by 2026 and 15 percent (228 million) by 2036 (Technical Group on Population Projections, 2019). The extent of population aging and the pace at which aging occurs would substantially differ across the states and population groups, as seen from the census and survey information and from the projections (Office of the Registrar General and Census Commissioner, 2011; 2019; Various rounds of National Family Health Survey). Table 1 provides information on population aging in India since 1981.

**Table 1.** Progress in Population Aging in India 1981-2036.

| Year | Number of persons aged 60 and<br>above (Million) | Percent of the total<br>population | Old Age<br>Dependency Ratio |
| --- | --- | --- | --- |
| 1981 | 43.1 | 6.3 | 12.0 |
| 1991 | 56.6 | 6.8 | 12.2 |
| 2001 | 76.6 | 7.5 | 13.0 |
| 2011 | 101.5 | 8.4 | 13.8 |
| 2021 | 137.9 | 10.1 | 15.7 |
| 2026 | 163.2 | 11.5 | 17.6 |
| 2031 | 193.8 | 13.1 | 20.1 |
| 2036 | 228.0 | 15.0 | 23.1 |

Source: Data for 1981-2011 are based on Census data. The projections made by the Registrar General and Census Commissioner (2019) are used for the remaining years.

Using census data and population projections, researchers have already observed that population aging in India occurs and progresses differently across states and population groups (Bhagat & Kumar, 2011; Dhar Chakrabarti, 2001; Dommaraju, 2016). There are also interstate differences in population aging; data from the Census of India 2011 indicate that the share of the older population varies from 7 percent in Manipur to 12.6 percent in Kerala. Among Indian states, the top states with a high share of the older population are Kerala (12.6 percent), Goa (11.2 percent), Tamil Nadu (10.4 percent), Punjab (10.3 percent), Himachal Pradesh (10.2 percent) and Maharashtra (9.9 percent). However, since the population size varies considerably across Indian states, the actual number of older populations would be different across states. For instance, Uttar Pradesh, which is the most populated state in India and had a population of 200 million in 2011, has 7.7 percent of its population aged 60 and above; translated into absolute numbers, this would be 15.4 million persons compared to Kerala, which has the highest proportion of older population with an absolute number of 4.2 million of older population. Depending on the developmental situation in different states, societal attributes and priorities, and policy and program arrangements, each state will have to make provisions to ensure the well-being of the increasing number of older persons.

Table 2 provides estimates of gender and rural-urban differences in population aging in 2021. To obtain this indication, we used the projected total population and age distribution of India’s population for 2021, the projected urban population for India 2021, and the age distribution of rural and urban populations as given in the NFHS 5 report. Using this information, Table 2 presents the estimated percent and the number of older persons in India in 2021. According to this, while in urban areas, the proportion of the older population is the same (1.6 percent) for males and females, the proportion is lower for females (11.4 percent) than for males (12.4 percent) in rural areas. In India, the estimated number of older individuals is 84.6 million males and 76.2 million females. However, if we use the projections used by the Census of India, the corresponding numbers are slightly lower: 67.1 million and 70.9 million, respectively (the percentage of those aged 60 and above according to census projections is 9.6 among males and 10.7 among females).

**Table 2.**
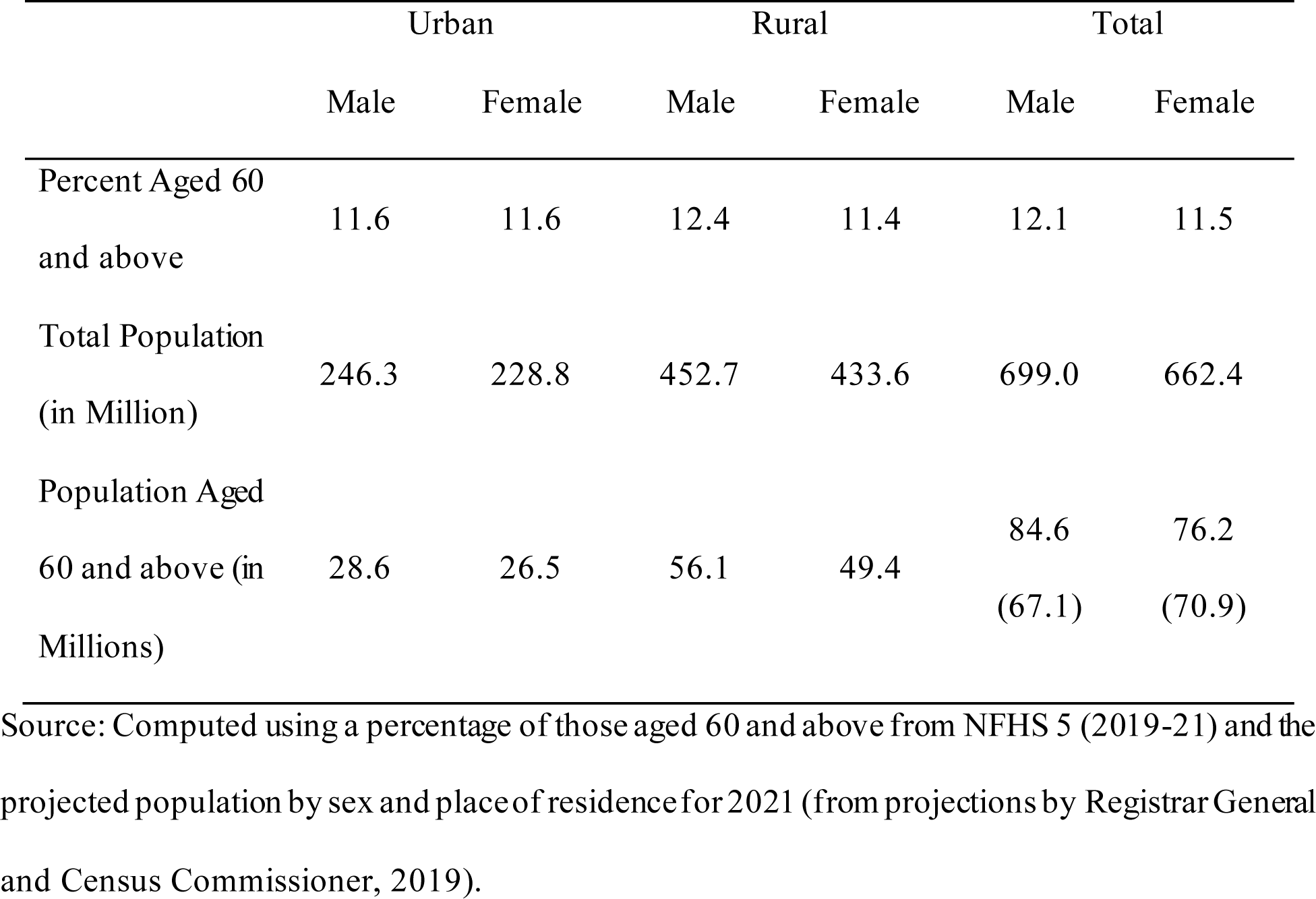
Percentage and number of persons aged 60 and above, according to sex and place of residence, India 2021.

|  | Urban |  | Rural |  | Total |  |
| --- | --- | --- | --- | --- | --- | --- |
|  | Male | Female | Male | Female | Male | Female |
| Percent Aged 60 and above | 11.6 | 11.6 | 12.4 | 11.4 | 12.1 | 11.5 |
| Total Population (in Million) | 246.3 | 228.8 | 452.7 | 433.6 | 699.0 | 662.4 |
| Population Aged 60 and above (in Millions) | 28.6 | 26.5 | 56.1 | 49.4 | 84.6<br>(67.1) | 76.2<br>(70.9) |

Source: Computed using a percentage of those aged 60 and above from NFHS 5 (2019-21) and the projected population by sex and place of residence for 2021 (from projections by Registrar General and Census Commissioner, 2019).

## 3. Impact of Population Aging on Health

Nearly one-fifth of the world’s population will be aged 60 and up by 2050 (United Nations, 2017). In India, the share of the older population is expected to be 20 percent from 8 percent in 2015 to 2050 (UNFPA, 2017). However, this shift presents the advanced need for more proper planning and strategies for present and upcoming problems related to healthy aging (Adlakha et al., 2020). SDG Goal 3, “Good health and well-being,” and WHO’s concern on “Healthy aging” on the line of health and well-being, it becomes crucial to understand the country’s aging structure, health profile, and available facilities. (WHO, 2015).

India, a developing country and currently the most populated country, has been experiencing an unpredictable increase in the proportion of the older population aged 60 and up. In 2014, the growth rate of the older population was three times higher than that of the overall population (Arokiasamy, 2016). Along with the unprecedented aging population, the health profile is also changing (Agarwal et al., 2016). In India, 95 percent of older persons suffer from at least one disease (Karmakar et al., 2014; Naushad et al., 2016). NCDs are major diseases among older populations, and this disease burden has also increased over the years. (Devadasan, 2006; Registrar general of india, 2013; Yadav & Arokiasamy, 2014). In 2016, Jayakrishnan, based on the NSSO survey conducted from January to June 2014, mentioned that the proportion of ailing older persons has increased over the last ten years (Jayakrishnan, 2016).

India has been experiencing an increase in urban population and is expected to have 40 percent of the people in urban areas by 2030; issues of easy accessibility and affordability are also increasing (Rao & Peters, 2015). While most large cities make it possible to manage good facilities, many smaller cities are still struggling. A difference in terms of development indicators and facilities was observed among the cities (Goli et al., 2011). Rao and Peters stated that a higher concentration of healthcare providers is in comparatively higher urbanized areas, yet not every urban area has easy access to health facilities (Rao & Peters, 2015). Most advanced health diagnostic systems and treatment facilities are concentrated in major cities and have different health systems and delivery structures (Agarwal et al., 2009). Therefore, one may expect a difference between large and small cities in terms of health facilities and accessibility.

However, the growth rate or proportion of the older population in the cities was almost the same (Census of India, 2011). It should be noted that smaller cities also have the same proportion of the older population and older adults suffering from age-related health problems; it becomes necessary to understand that these cities struggle to obtain health coverage at the right time. This situation drives wide-ranging challenges to cities and the government in maintaining health coverage, especially for the older population. India also launched a National Urban Health Mission (NUHM) program in 2013 with the prime objective of “healthy cities with good health facilities.” However, in India, most urban planners currently overlook the necessity of making age-friendly cities (Adlakha et al., 2020).

While there has been considerable improvement in health facilities over the years and across cities, if we look at smaller cities, health facilities are not as improved as in large cities. People from smaller cities have to travel great distances to receive quality treatment. With the equal proliferation of health facilities and coverage, the gap within cities needs to be examined. Previous literature was engaged to raise the problems between rural and urban, slum and non-slum, and major cities. Nevertheless, hardly any studies discuss differences between health facilities and other development indicators among the different cities (Garg & Karan, 2009; Zare et al., 2018). Thus, the present study aimed to examine the distribution of the older population in Indian cities. Additionally, the study tried to investigate access to improved health facilities among different types of cities in India.

## 4. Methods

While there are recent national-level surveys in India, none of them provide data for all cities in India. Thus, the study used data from the Census of India (2011) which is the latest census of India. The census provides age-groupwise population data for Class I cities. Additionally, data on health facilities were collected from the Town Directory data of 2011. The town directory provides data on available health facilities in a city. Data on older populations above age 60 were also collected from the Census C-14 (City) table for 2001 and 2011. A total of 492 cities were studied in this research. The share of the older population was calculated by dividing the share of older adults (60 years and above) by the total population. Additionally, we have also estimated the share of the older population for the year 2021 using the exponential growth method.

To understand the available health facilities in Class I cities, a Health Facilities Index (HFI) score was calculated. Available health facilities, such as the number of hospitals, doctors, paramedical staff, and hospital beds, were taken into consideration, and a final composite score was computed. The mean distance to these health facilities was also calculated from the dataset. Equal weightage was given to all the indicators to prepare the index. Furthermore, the variation in the share of the older population has been understood by the size and status of the cities. Similarly, available health facilities were compared with the aging scenario in these cities by size and status.

ArcGIS software was used to map the share of the older population in Indian cities. We have tried to project the share of the older population for 2021. An inventory map was created to present the available health facilities in a city and the average distance to the health facility. Hotspot analysis was also performed to identify the major hotspots of aging in India. The following table shows the indicators and sub-indicators used to compute the health facility index.

**Table 3:** List of indicators and sub-indicators of the Health Facilities Index.

| Indicators of Health Facilities | Sub-indicators used (Per 000' Population) |
| --- | --- |
| 1. Government Hospital | a. Hospital Allopathic Numbers |
| 2. Allopathic Government Hospital Alternative | b. Number of Beds |
| 3. Dispensary/Health Centre | c. Number of Doctors |
| 4. T.B Hospital/Clinic | d. Number of Para Medical Staffs |
| 5. Nursing Home |  |
| 6. Mobile Health Clinic |  |
| 7. Others |  |

## 5. Results

### 5.1. Aging pattern in Indian cities

Figure 1 shows the distribution of older populations among the different city sizes. The older population was 4.7 million in million-plus cities, which doubled (10.3 million) in 2011. Other cities had a smaller share of older people compared to million-plus cities. Cities with less than 2 lakh population have 2.9 million, cities with 2-5 lakh population have 3.5 million, and those with 5-10 lakh population have 2.5 million older populations. This increase in the number of older people was rapid in million-plus cities but slightly lower in other smaller cities during 2001-11.

**Figure 1:**
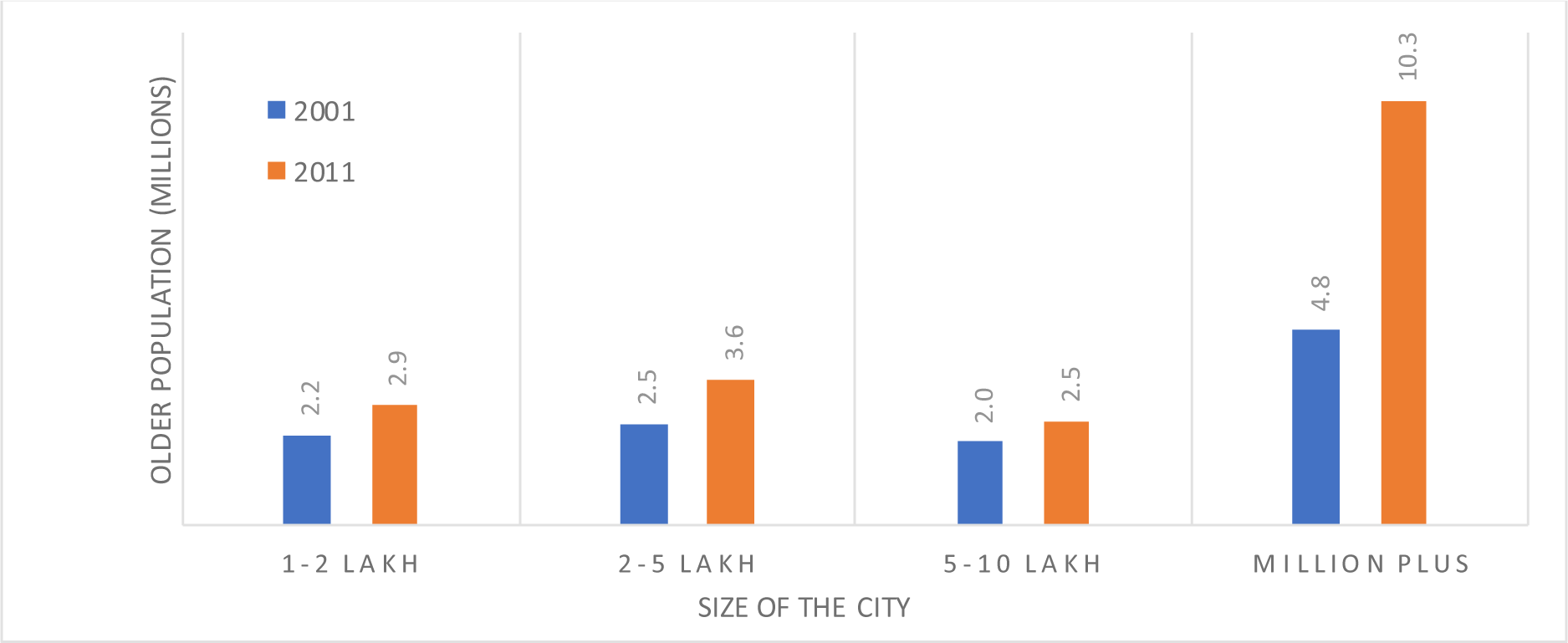
Older population by city size in 2001 and 2011.

The share of the older populations (60 years and above) increased among different sizes of cities during 2001-11. The highest increase was observed among the cities with 5-10 lakhs of the population between 2001 and 2011. Here, it is observed that cities with smaller populations have a slightly higher share of older people than larger cities. Cities with a population of less than a million have a population of 8.2 percent, while million-plus cities have a population of 7.9 percent older. The study also highlights that there is a wide variation in the distribution of older populations across cities with various civic statuses. Here, census towns have 5.5 percent of the older population, the lowest, and municipalities have the highest share, which is 9.3 percent. An increase in the percentage of the older population can be observed across all types of cities during 2001-11 (Table 4).

**Table 4:**
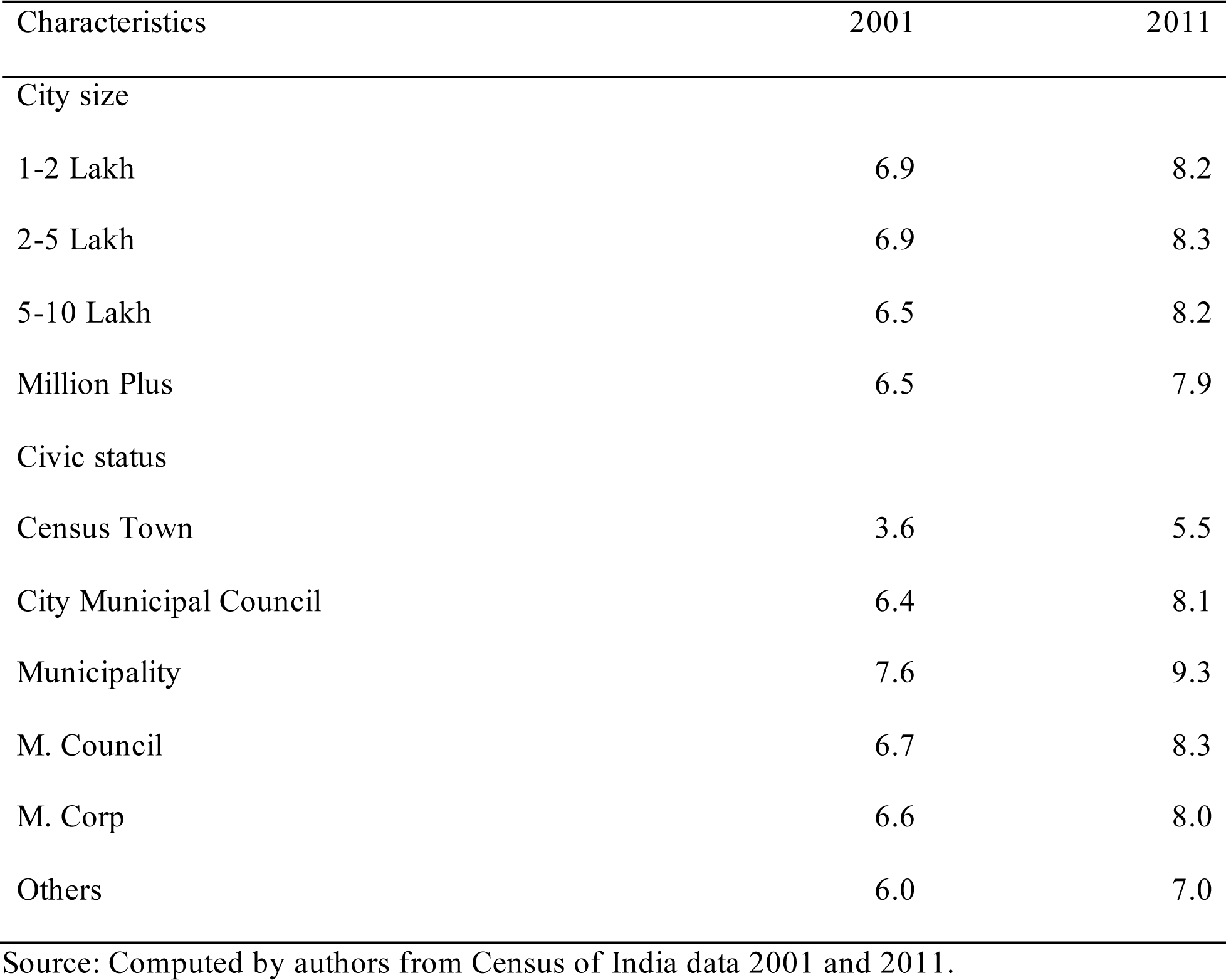
Share of older population by city size and civic status in India 2001-11.

Regional variations in the distribution of the older population among Class I cities are presented in Figure 2. In 2001, most of the cities had an older population of less than 8 percent, but states such as Kerala, Tamil Nadu, and West Bengal had a higher share of the older population (more than 10 percent). A rapid increase can be observed during 2001-2011 across the states and cities.

**Figure 2:**
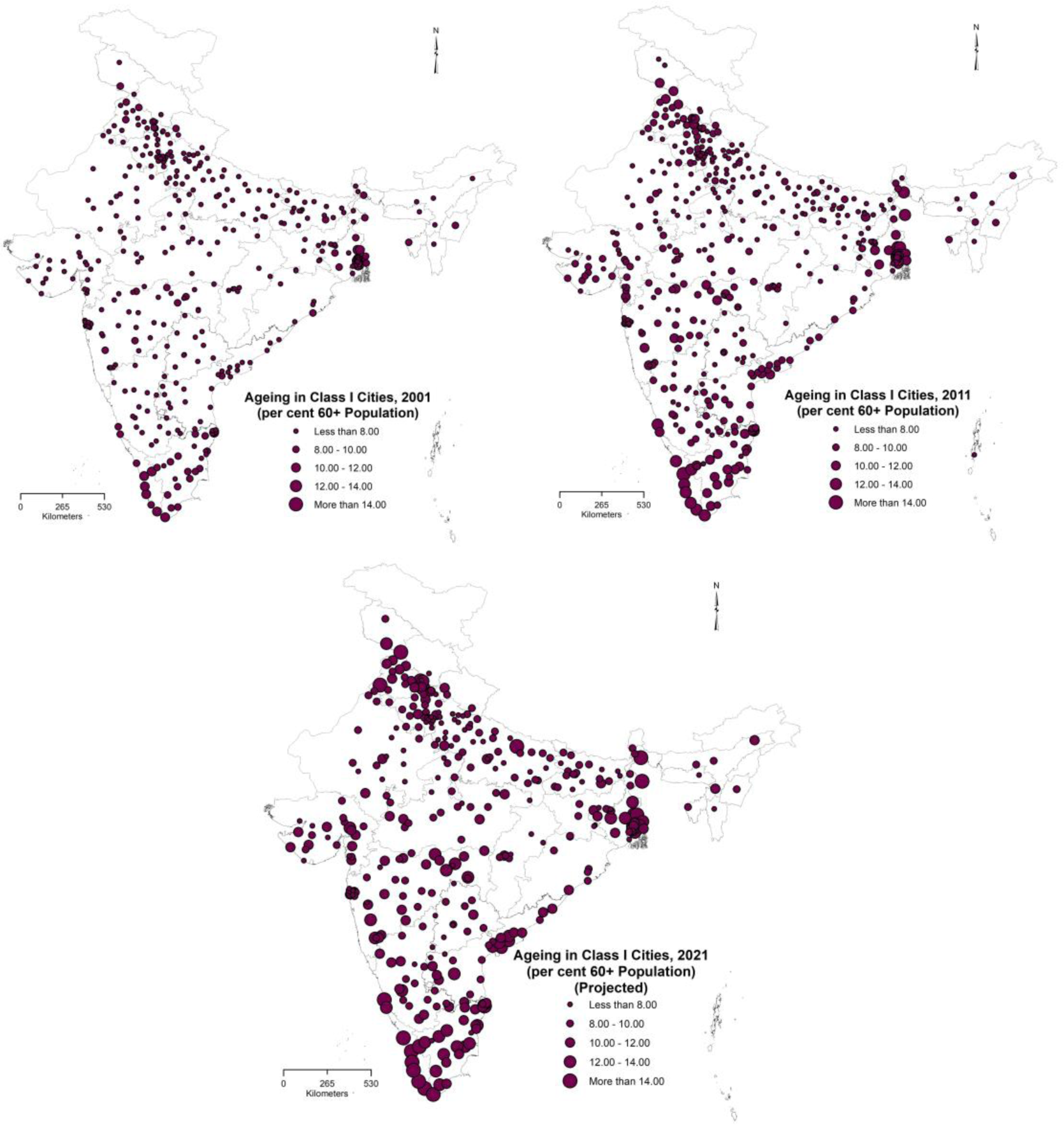
Distribution of the older population (60 years and above) among Class I cities for 2001, 2011, and 2021 (Predicted) in India.

The concentration of cities with a higher share of the older population can be found in states such as Kerala, Tamil Nadu, and West Bengal. The percentage of the older population is expected to increase above 14 percent in many cities across the country. The major concentration of the older population will be observed in Kerala, Tamil Nadu, West Bengal, Punjab, Delhi, Maharashtra, etc. Thiruvananthapuram, Palakkad, Hugli-Chinsurah, Khardah, South DumDum, Uttarpara Kotrung, and Nabadwip.

Hotspot analysis shows the hotspots for the older population in India (Figure 3). The southernmost and eastern parts of India are the two major hotspots with 99 percent confidence. Most cities have a higher proportion of the older population and are surrounded by cities with the same. The cold spot is the northern part, consisting of Punjab, Haryana, Uttar Pradesh and Rajasthan. The share of the older population in the central part of India is not clustered; therefore, it is insignificant.

**Figure 3:**
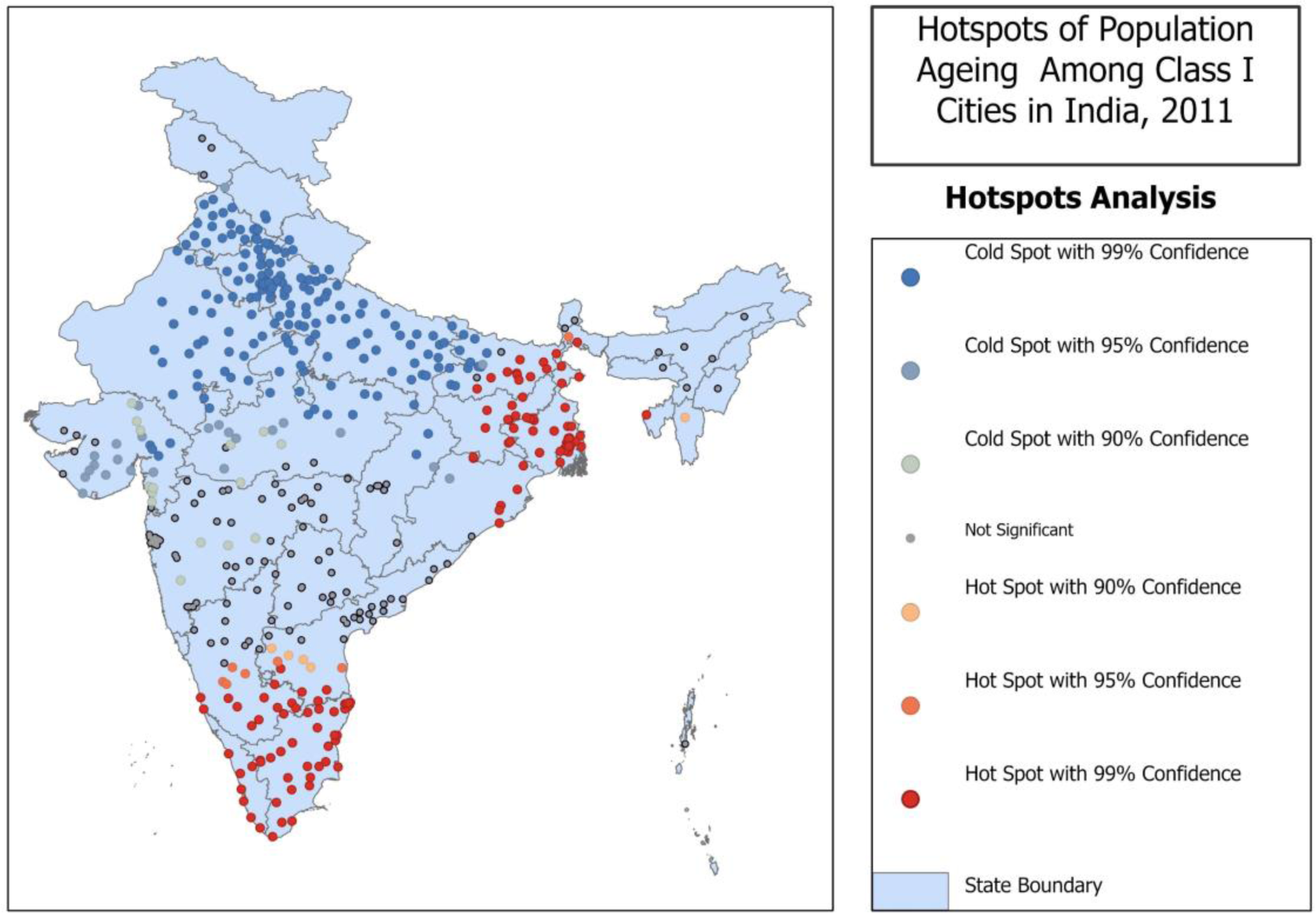
Hotspots of older populations among Class I cities in India, 2011

### 5.2. Aging and health facilities in Indian cities

Health facilities are primarily associated with population size among the cities in India. Figure 4 shows that cities with larger populations have better and more health facilities. In comparison, cities with smaller populations have a smaller number of improved health facilities, as evidenced by the HFI scores.

**Figure 4:**
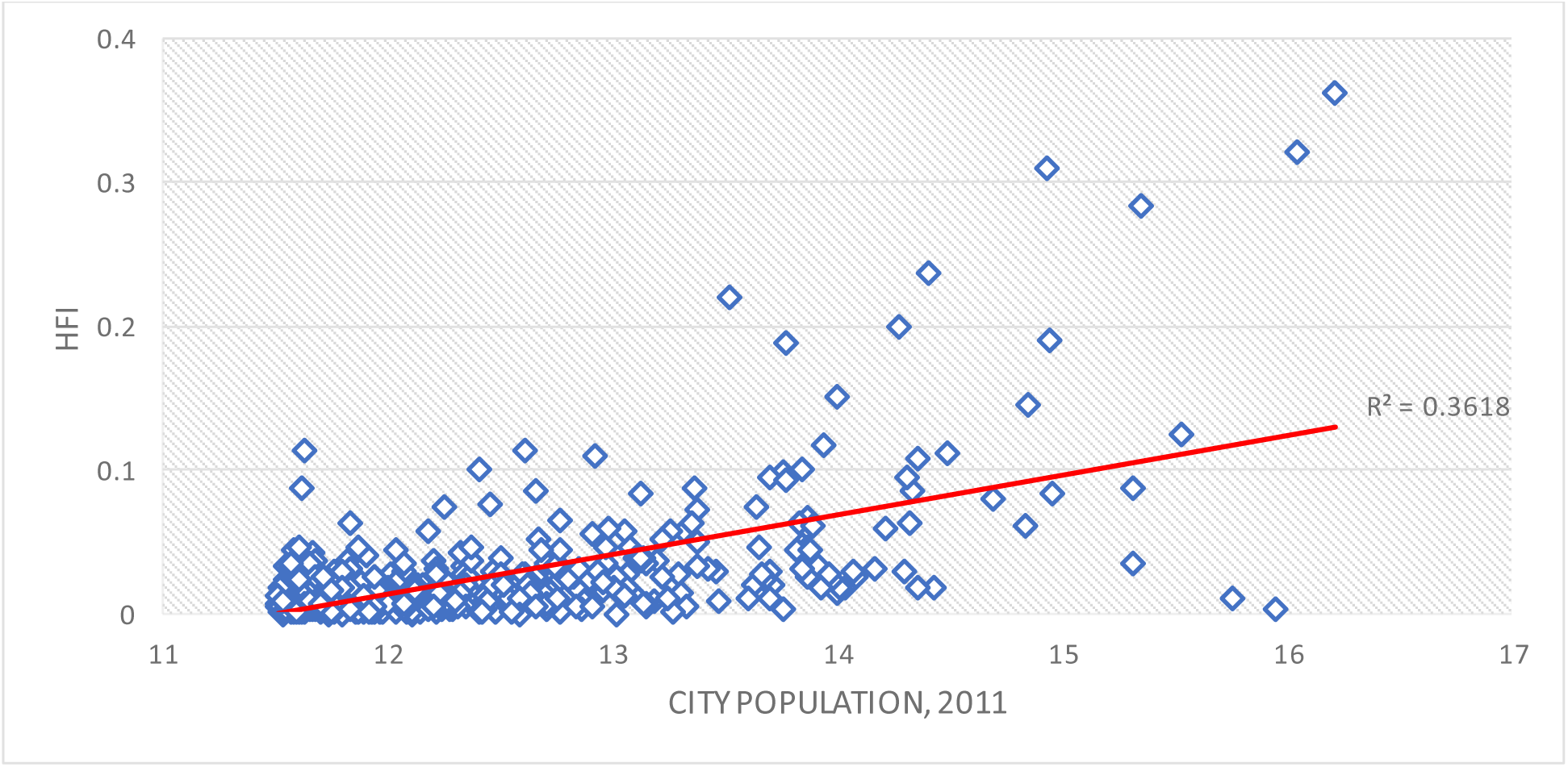
The relationship between the population size of cities and the HFI score, 2011

There is a gap between the proportion of older population and health facilities across different-sized Class I cities. Table 4 shows that the share of the older population is almost the same in cities of different sizes, but health facilities are not the same. Although the million-plus cities have a slightly lower percentage of the older population, they have comparatively better health facilities. At the same time, other cities have fewer health facilities even though the share of the older population is higher.

**Table 4:** Comparison of the older population share and mean Health Facilities Index, 2011.

| Characteristics | Share of older Population | Mean HFI |
| --- | --- | --- |
| City size |  |  |
| 1-2 lakh | 8.2 | 0.01 |
| 2-5 lakh | 8.3 | 0.02 |
| 5-10 lakh | 8.2 | 0.04 |
| Million Plus | 7.9 | 0.09 |
| Civic status |  |  |
| CT | 5.5 | 0.01 |
| NP | 6.8 | 0.01 |
| CMC | 8.1 | 0.02 |
| M | 9.3 | 0.01 |
| MCI | 8.3 | 0.02 |
| Mcorp | 8.0 | 0.1 |
| Others | 7.7 | 0.03 |

Source: Computed by authors from Census of India data 2001 and 2011

There is variation in the share of the older population and available health facilities among cities with different civic statuses (Table 4). Municipal Corporation has better health facilities among different civic bodies, with an eight percent share of older populations. The HFI score is very low in census towns, Nagar panchayats, and municipal councils, even though these cities have a decent share of the older population. Municipalities have the highest share of older people (9.3 percent), but the HFI score is very low (0.01).

Figure 5 portrays the insufficiency of health facilities with respect to population aging in cities in India. It clearly shows that only a few major cities, such as Jaipur, Bangalore, Chennai, Mumbai, Pune, Kolkata, and Delhi, have a sufficient level of health facilities with respect to the older population. At the same time, many cities that have experienced rapid population aging in the last decade lack health facilities. Emerging aging cities located in states such as Kerala, Tamil Nadu, and West Bengal are very much lacking in health facilities where the older population is already higher. Therefore, improving health facilities in these cities is an urgent need of the time for healthy urban aging.

**Figure 5:**
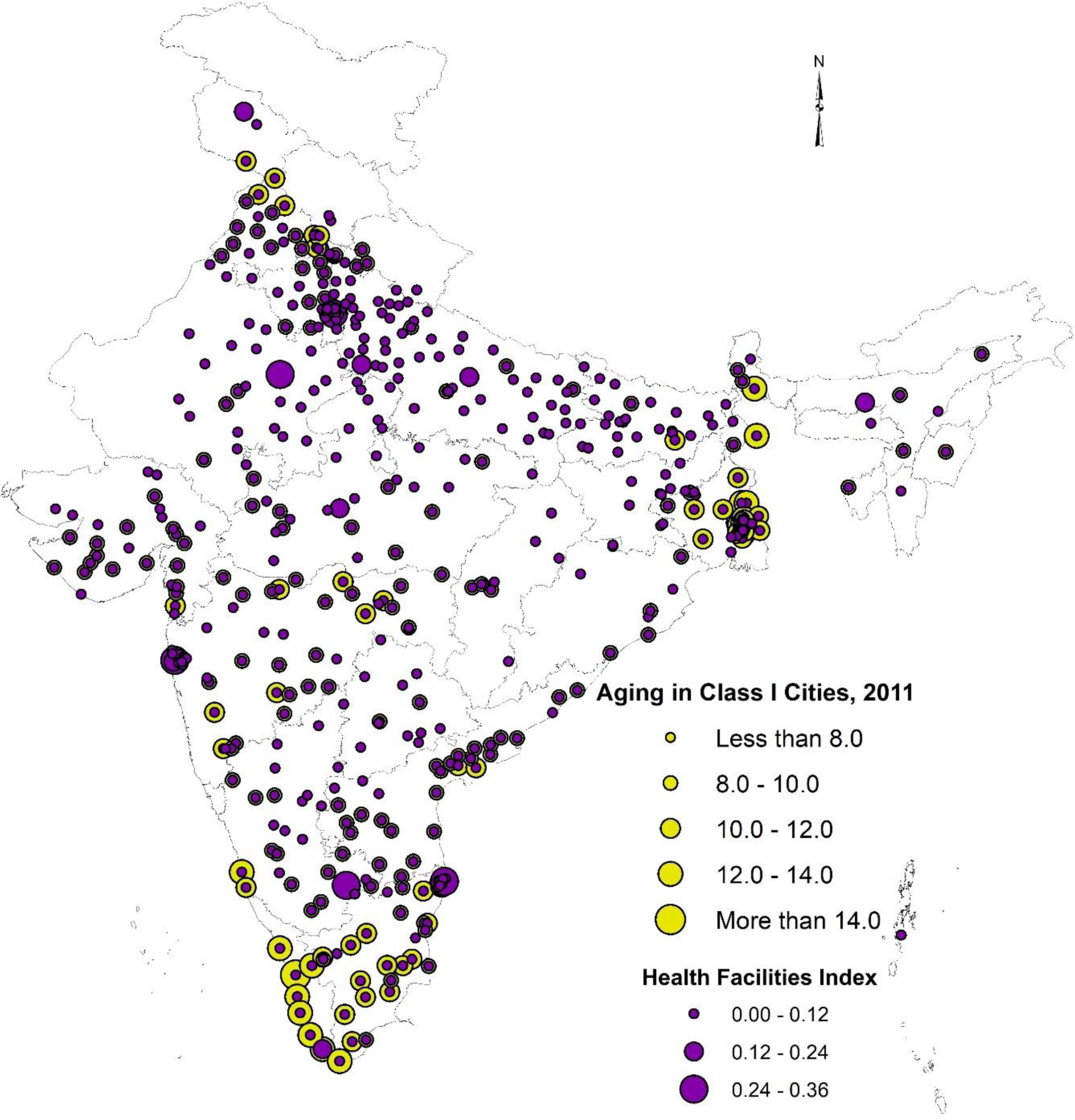
Share of older population and available health facilities in Class I cities in India, 2011.

The examination of the health facility infrastructure across various cities in India has brought to light some fascinating findings (Figure 6). In particular, we noticed that larger cities such as Bengaluru, DMC, Mumbai, Jaipur, Chennai, Bhopal, Thiruvananthapuram, Agra, and Guwahati boast high health facility scores. This is a promising indicator, as it suggests that these cities have the requisite resources and infrastructure to cater to the health needs of their population, especially older adults. The study shows that cities with lower health facility scores have a significantly longer distance to travel to access health facilities. This is especially true for cities in Uttar Pradesh, Bihar, and some cities in Madhya Pradesh, Maharashtra, and Gujarat. In contrast, cities in Kerala and Tamil Nadu have more accessible health facilities, with relatively shorter distances to travel.

**Figure 6:**
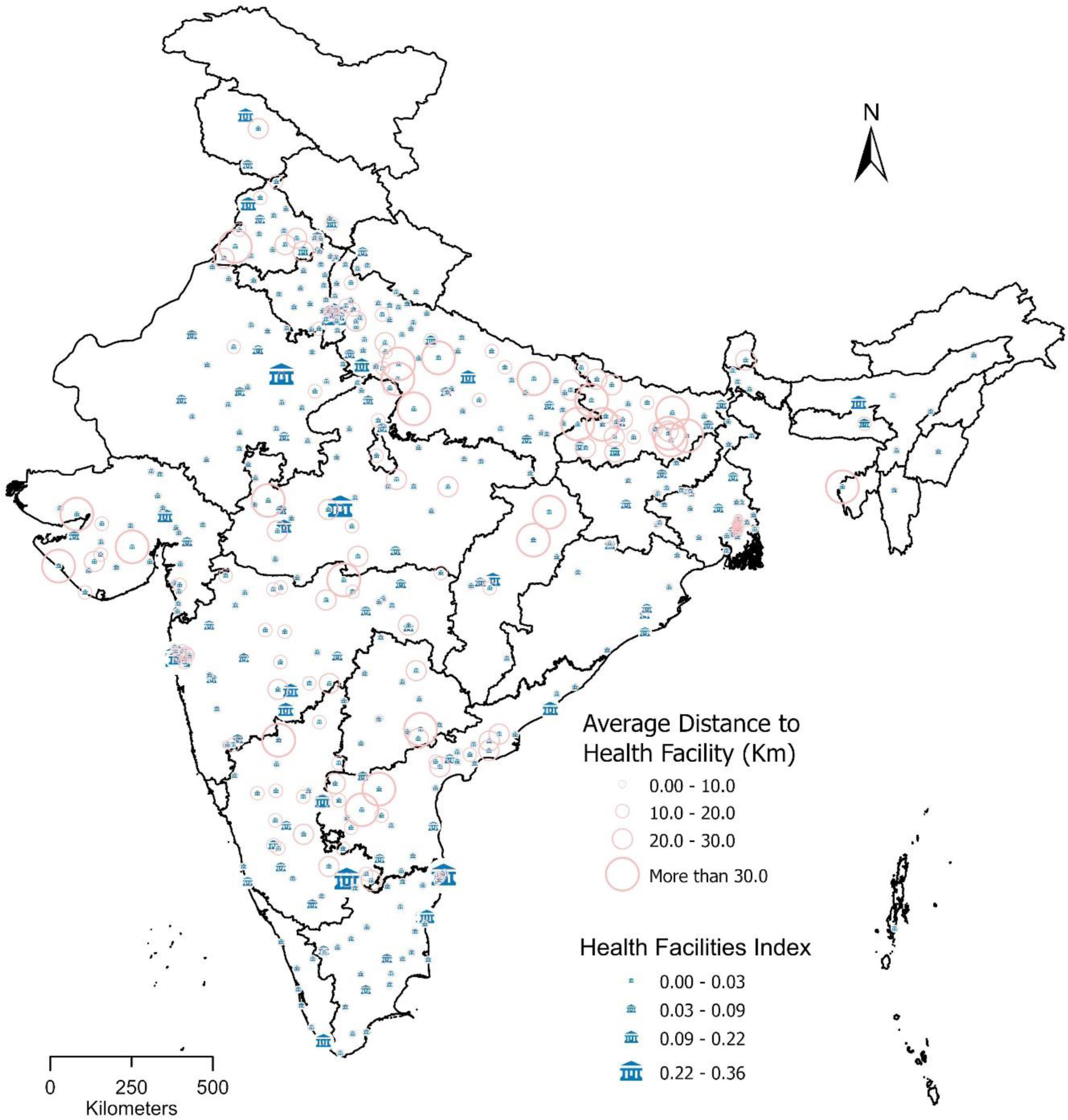
Existing health facilities and average distance to the health facility among Class I cities in India, 2011.

## 6. Discussion

This study aimed to investigate the relationship between population aging and available health facilities in Indian cities. While many studies have focused on the aging population at the state level, few have examined the situation at the city level. It was found that Indian cities have a significant share of the older population, and the numbers have increased rapidly in the last decade. However, there are considerable differences in health facilities across regions, and many cities suffer from limited health infrastructure and staff. The study compared the distribution of older adult populations and health infrastructures, such as hospitals and medical staff, and found considerable regional variations in their availability.

Smaller cities tend to have poorer health facilities than larger cities, despite having similar proportions of older populations. As India’s demographic structure changes, it is essential to plan and prepare for the needs of the aging population (United Nations, 2020). The study highlights the need for policies and programs targeting the older population and for extensive research on changing population aging patterns in cities. Urban planning must ensure the creation of aging-friendly environments to meet the needs of the older population.

With the aging of the increasing population, it is crucial to adequately plan for the health and well-being of older people (Gardener & Lemes de Oliveira, 2020; Noroozian, 2012). However, there are significant variations in the availability of health facilities across different regions in India. While urbanization is often seen as a marker of development, it is important to note that different urban areas are at varying stages of development. While previous studies have focused on the lack of healthcare facilities in rural areas, there is a pressing need to investigate the extent of healthcare facilities in urban India, specifically in cities. Therefore, the study aimed to understand the level of accessibility and availability of advanced healthcare facilities in various cities.

In this study, we examined the distribution of the older population and the available health infrastructure, such as hospitals, hospital beds, and healthcare personnel, in various cities. We found that a significant number of cities in India have inadequate health infrastructure, staff, and facilities (Ajitha et al., 2022). Moreover, there are regional differences in the distribution of older populations among Class I cities, with a higher concentration in the northern, southern, and eastern parts of India.

Cities with a larger population generally have better-advanced health facilities, while smaller cities suffer from a lack of good health facilities, even though they have similar proportions of older populations. Therefore, it is crucial to focus on the health infrastructure of smaller cities as much as larger ones. Our study also revealed that there are significant variations in infrastructure, facilities, and services among cities, as well as in travel distance to health facilities (Bhan et al., 2017; Hoof et al., 2018). Additionally, the number of programs aimed at the older population is limited and requires updating in India.

While the study has provided essential information regarding the adequacy of health facilities for an aging population in India, it is not without limitations. The study has used data from the Census of India, 2011 as the new census is yet to take place. Thus, due to the relatively old data, the estimates may differ from the actual scenario.

## 7. Conclusions

This study looks at the distribution patterns of the aging population as well as the inadequacies affecting healthcare infrastructure, which includes not only hospitals and hospital beds but also vital healthcare personnel, across a wide range of cities of varying size and classification. The study’s findings emphasize the evident shortage of basic health facilities, competent staff, and critical resources throughout multiple cities dispersed over India.

Furthermore, the disparities that exist on a regional level that concern the distribution of the older population among the cities categorized as Class I are impossible to overlook. While the larger urban centers inherently enjoy an advantage in terms of possessing well-equipped, advanced health facilities, it is very different in the case of their smaller counterparts. Despite similar proportions of older people, the smaller cities face persisting inadequacies, emphasizing a discrepancy that needs to be looked into immediately.

Given these significant findings, we must focus on and enhance the quality of healthcare services in smaller cities. This implies that we must ensure that smaller cities have the same resources and capacities as larger cities. The absence of appropriate healthcare alternatives is a major issue th at impacts the whole Indian healthcare system. It is necessary to think broadly and prepare for the changes that will occur as more people age in the future. This means that we should have a comprehensive strategy in place to assist individuals in aging in a healthy manner.

Hence, it is imperative to prioritize the enhancement of health infrastructure in smaller cities on par with larger ones. The inadequacy of health infrastructure emerges as a major issue in the Indian healthcare system. However, it is crucial to prioritize preparedness for forthcoming demographic changes in the country. This concept of assisting individuals in ageing properly might be the key to long-term success. It may also create new opportunities for the economy to flourish. By achieving healthy aging, we can unlock the door to productive aging and seize the economic opportunities that lie ahead.

## Data availability statement

The present study used secondary data collected from the Census of India, 2011. The town directory data are freely available for use. It can be downloaded from the following website.

https://censusindia.gov.in

## References

Adlakha, D., Krishna, M., Woolrych, R., & Ellis, G. (2020). Neighbourhood Supports for Active Ageing in Urban India. Psychology and Developing Societies, 32(2), 254–277. 10.1177/0971333620937497

Agarwal, A., Lubet, A., Mitgang, E., Mohanty, S., & Bloom, D. E. (2016). Population Aging in India: Facts, Issues, and Options. SSRN Electronic Journal, 10162. 10.2139/ssrn.2834212

Agarwal, S., Health, U., Centre, R., Srivastava, A., Kaushik, S., & Banerjee, P. (2009). Diversity in health challenges across cities in India: Need for context specific response. Journal of Urban Health, 86(3), 389–497. 10.1007/s11524-009-9361-8

Ajitha, D., Gouri, C. S., Eklure, S. B., & Chakraborty, C. (2022). Healthcare Infrastructure in Future Smart Cities. In Intelligent Healthcare (pp. 321–341). 10.1007/978-981-16-8150-9_15

Arokiasamy, P. (2016). Population Ageing in India. Ageing and Society, 36(2), 445–447. 10.1017/s0144686x15001300

Bhagat, R. B., & Kumar, K. (2011). Ageing in India: Trends and Patterns. January 2011, 19–37.

Bhan, N., Madhira, P., Muralidharan, A., Kulkarni, B., Murthy, G., Basu, S., & Kinra, S. (2017). Health needs, access to healthcare, and perceptions of ageing in an urbanizing community in India: A qualitative study. BMC Geriatrics, 17(1), 1–11. 10.1186/s12877-017-0544-y

Census of India. (2011). Census of India 2011 Meta Data. In Office of the Registrar General & Census Commissioner, India. https://censusindia.gov.in/census.website/data/census-tables

Devadasan, N. (2006). Health financing : Protecting the poor. Indian Journal of Community Medicine. 10.4103/0970-0218.54922

Dhar Chakrabarti, P. G. (2001). Urban crisis in India: New initiatives for sustanable cities. Development in Practice, 11(2–3), 260–272. 10.1080/09614520120056397

Dommaraju, P. (2016). Contemporary Demographic Transformations in China, India and Indonesia. Contemporary Demographic Transformations in China, India and Indonesia, January 2016, 0–21. 10.1007/978-3-319-24783-0

Gardener, M. A., & Lemes de Oliveira, F. (2020). Urban environment cues for health and well-being in the elderly. Cities and Health, 4(1), 117–134. 10.1080/23748834.2019.1636506

Garg, C. C., & Karan, A. K. (2009). Reducing out-of-pocket expenditures to reduce poverty: A disaggregated analysis at rural-urban and state level in India. Health Policy and Planning, 24(2), 116–128. 10.1093/heapol/czn046

Goli, S., Arokiasamy, P., & Chattopadhayay, A. (2011). Living and health conditions of selected cities in India: Setting priorities for the National Urban Health Mission. Cities, 28(5), 461–469. 10.1016/j.cities.2011.05.006

Hoof, J. van, Kazak, J. K., Perek-Białas, J. M., & Peek, S. T. M. (2018). The challenges of urban ageing: Making cities age-friendly in Europe. International Journal of Environmental Research and Public Health, 15(11), 1–17. 10.3390/ijerph15112473

International Institute for Population Sciences. (2020). National Family Health Survey - 5. In Ministry of Health and Family Welfare National (Vol. 361).

Jayakrishnan, T. (2016). Increasing Out-Of-Pocket Health Care Expenditure in India-Due to Supply or Demand? Pharmacoeconomics, 01(01). 10.4172/2472-1042.1000105

Karmakar, P. R., Chattopadhyay, A., & Sarkar, G. N. (2014). A Study on Morbidity Pattern and Care Seeking behaviour of Elderly in a Rural Area of West Bengal (India). Indian Journal of Gerontology, 28(2), 190–200.

Naushad, M. A., Verma, N., Bhawnani, D., Jain, M., Amand, T., & Umate, L. V. (2016). Morbidity pattern and health seeking behavior in elderly population of Raipur City, Chhattisgarh, India. International Journal for Equity in Health, 28(03), 236–241.

Noroozian, M. (2012). The elderly population in iran: An ever growing concern in the health system. Iranian Journal of Psychiatry and Behavioral Sciences, 6(2), 1–6.

Rao, K. D., & Peters, D. H. (2015). Urban health in India: Many challenges, few solutions. The Lancet Global Health, 3(12), e729–e730. 10.1016/S2214-109X(15)00210-7

Registrar general of india. (2013). Report on Medical Certification of Report on Medical Certification of.

Rele, J. R. (1987). Fertility Levels and Trends in India, 1951-81. 13(3), 513–530.

Technical Group on Population Projections. (2019). Population projections for India and states 2011-2036. https://nhm.gov.in/New_Updates_2018/Report_Population_Projection_2019.pdf

UNFPA. (2017). Caring for Our Elders : Early Responses India Ageing Report-2017. United Nations Population Fund, 33(1), 531–540.

United Nations. (2017). World Population Ageing 2017. In World Population Ageing 2017. http://www.un.org/en/development/desa/population/publications/pdf/ageing/WPA2017_Report.pdf

United Nations. (2019). World population prospects 2019. In Department of Economic and Social Affairs. World Population Prospects 2019.

United Nations. (2020). World Population Ageing. In Economic and Social Affairs United Nations. http://link.springer.com/chapter/10.1007/978-94-007-5204-7_6

United Nations. (2022). World Population Prospects. In World Population Prospects. 10.18356/cd7acf62-en

WHO. (2015). World Report On Aging and Health.

Yadav, S., & Arokiasamy, P. (2014). Understanding epidemiological transition in India. Global Health Action, 7(SUPP.1). 10.3402/gha.v7.23248

Zare, V. R., Kokiwar, P., & Ramesh, B. (2018). Health status of elderly: a comparative study among urban and rural dwellers. International Journal Of Community Medicine And Public Health, 5(7), 3039. 10.18203/2394-6040.ijcmph20182645

